# Effect of hospital assignment on mortality for AMI patients

**DOI:** 10.1101/2022.07.08.22277206

**Authors:** Mari Grøsland, Kjetil Telle, Henning Øien

## Abstract

International guidelines recommend percutaneous coronary intervention (PCI) to treat acute myocardial infarction (AMI) if PCI can be performed within two hours. PCI is a centralized treatment, and therefore a common trade-off is whether to send AMI patients directly to a hospital that performs PCI, or postponing a potential PCI-treatment by first receiving acute treatment at a non-PCI-hospital. In this paper, we estimate the effect of sending patients directly to a PCI-hospital on AMI mortality. Since the underlying health of patients may affect both hospital assignment and mortality, estimates from traditional multivariate risk adjustment models are likely biased. We therefore apply an instrumental variable (IV) model using the historical municipal share that is sent directly to a PCI-hospital as an instrument for being sent directly to a PCI-hospital. Our results disclose that patients sent directly to a PCI-hospital are younger and have fewer comorbidities than patients who are first sent to a non-PCI-hospital. Our IV results suggest that those initially sent to PCI-hospitals have 4.8 percentage points *decrease* (95% CI (-18.1)-8.5) in mortality after one month compared to those initially sent to non-PCI-hospitals. However, the estimates are too imprecise to conclude that health personnel should change their practice.

## 1 Introduction

Acute myocardial infarction (AMI) is the leading cause of death in the industrialized world (OECD, 2019), and imposes large costs in terms of morbidity and reduced longevity, medical treatment, and work-related disabilities. Effective treatment of AMI is therefore essential to alleviate the individual and societal costs of the disease. The preferred emergency treatment of AMI is percutaneous coronary intervention (PCI) which is a non-invasive method that opens blocked coronary arteries. However, AMI is a time-critical condition – a successful outcome depends on treatment occurring quickly after onset of an attack – and PCI is a geographically centralized treatment. For example, in Norway only 8 of about 50 hospitals can perform PCI. If the patient is too frail or PCI cannot be performed within 2 hours after onset of the attack, thrombolysis (a drug that can dissolve the blood clot causing the blockage) is considered superior, at least in the short run. For patients living far away from a PCI-hospital there is therefore a trade-off between being sent directly to a PCI-hospital, or postponing a potential PCI-treatment by first receiving acute treatment at a local hospital.

In this paper, we explore the treatment effectiveness of being sent directly to a PCI-hospital for AMI-patients using observational data from Norway. The question is important to answer to provide evidence on the benefits of increasing the share sent directly to a PCI-hospital.There is, however, an endogeneity problem with the use of standard multivariate risk adjustment techniques when analyzing the importance of initially being sent to a PCI-hospital compared with being first admitted to a local non-PCI-hospital; the decision depends on health factors that is unobserved for researchers. The primary guidelines on treatment for AMI patients is therefore substantiated by randomized controlled treatment trials (RCTs) (Members et al., 2012). However, ethical, financial and practical issues complicate the use of such trials to answer the question of interest. In this paper, we illustrate this endogeneity problem by applying different analytical methods that to varying degrees takes this endogeneity problem into account. We first use multivariate risk adjustment methods, where we compare the health outcomes of patients who are sent directly to a PCI-hospital to patients who are first sent to a local non-PCI-hospital (and may or may not be transferred to a PCI-hospital later), while controlling for individual and demographic characteristics. We argue that it is likely that the treatment decision depends on unobserved health factors and employ an instrumental variable (IV) analysis to account for this. The instrument used is the historic share sent directly to a PCI-hospital in the patients’ municipality. Similar local area practice style instruments have been used in several earlier studies (Brooks et al. (2013); Stukel et al. (2007)).

Our results show as expected that patients sent directly to a PCI-hospital are younger and have fewer comorbidities compared to patients first sent to a local non-PCI-hospital. Multivariate risk adjustment methods suggest that those sent directly to a PCI-hospital have 1.4 percentage points (95% CI 0.84 to 1.96) higher mortality after 1 month than those sent to a non-PCI-hospital. However, using the historic municipal share that is sent to a PCI-hospital as an instrument, we find a negative point estimate of 4.8 percentage points (95% CI -18.1 to 8.5) in mortality. However, the estimates are too imprecise to conclude that health personnel should change their practice and send more patients directly to a PCI-hospital. Moreover, the results may be taken to suggest that health personnel navigate AMI patients to the best treatment option.

The rest of the article is organized as follows. In the next section, we explain how AMI is diagnosed, and describe the Norwegian setting. Then, in Section 3, we give a description of the data and methods. In Section 4, we present the results, and in the last section we discuss and make concluding remarks.

## 2 Background: Diagnosing AMI

AMI occurs when the blood supply to the heart is disrupted so that the heart does not get sufficient oxygen. Restoring blood flow to the affected part of the heart is therefore the primary therapeutic goal. This can be accomplished with medication or through invasive surgical procedures. If a heart attack is suspected, a medical expert should perform a echo-cardiogram (ECG) as soon as possible, which in many cases can be done before admission to hospital. The on-duty internal medicine physician on the local hospital should then be conferred with the results, and evaluate different treatment strategies. If the ECG shows an ST-elevation myocardial infarction (STEMI) the patient should, according to the guidelines from European Society of Cardiology (2012), be sent directly to a PCI-hospital if this is possible within 120 minutes. If this is not possible the patient should get thrombolysis, a medical treatment that can open the blockage, before the patient is sent to a PCI-hospital (Members et al., 2012).

The ECG, often in combination with a blood sample test, may also show that the patient have a non-ST-elevation myocardial infarction (nSTEMI). In 2011/2012 there was a significant reduction on the recommended time from first medical contact to evaluation at a PCI-hospital from within 24-78 hours, to within 2-78 hours, depending of the risk profile of the patient (*<*2 hours for patients with very high risk, *<*24 for those with high risk and *<*78 hours for the patients with lower risk profile) for these patients (Members et al. (2011); Wijns et al. (2010)).

Mortality after 30 days is higher for STEMI patients than nSTEMI patients, but after 6 months the mortality rates are very similar in both conditions (Marceau et al., 2013). Longer term follow-ups have shown that death rates were higher among patients with nSTEMI (Terkelsen et al., 2005), which may be explained by their higher incidence of co-morbidities such as kidney failure, diabetes and hypertension (van’t Hof and Badings, 2019).

Norway provides an attractive context to analyze effects of being admitted directly to PCI-hospitals for several reasons. First, Norway has universal health coverage, funded primarily through taxes and transfers from the national government. Thus, the treatment choice does not rely on health insurance coverage. Second, AMI is the leading cause of death in Norway, and detailed information on date of death for the whole population is recorded in national health registries. This gives us a well-defined endpoint. Third, suspected AMI is registered in the Norwegian Patient Registry (NPR), which is a complete patient registry containing information on all hospital treatments at the individual level since 2008. Additionally, many people live in rural areas but PCI-treatment is highly centralized, implying that health personnel must often decide to send an AMI patient to a local non-PCI-hospital or to a PCI-hospital farther away. This is seen in Figure 1 where we plot the location of PCI-hospitals.

**Figure 1:**
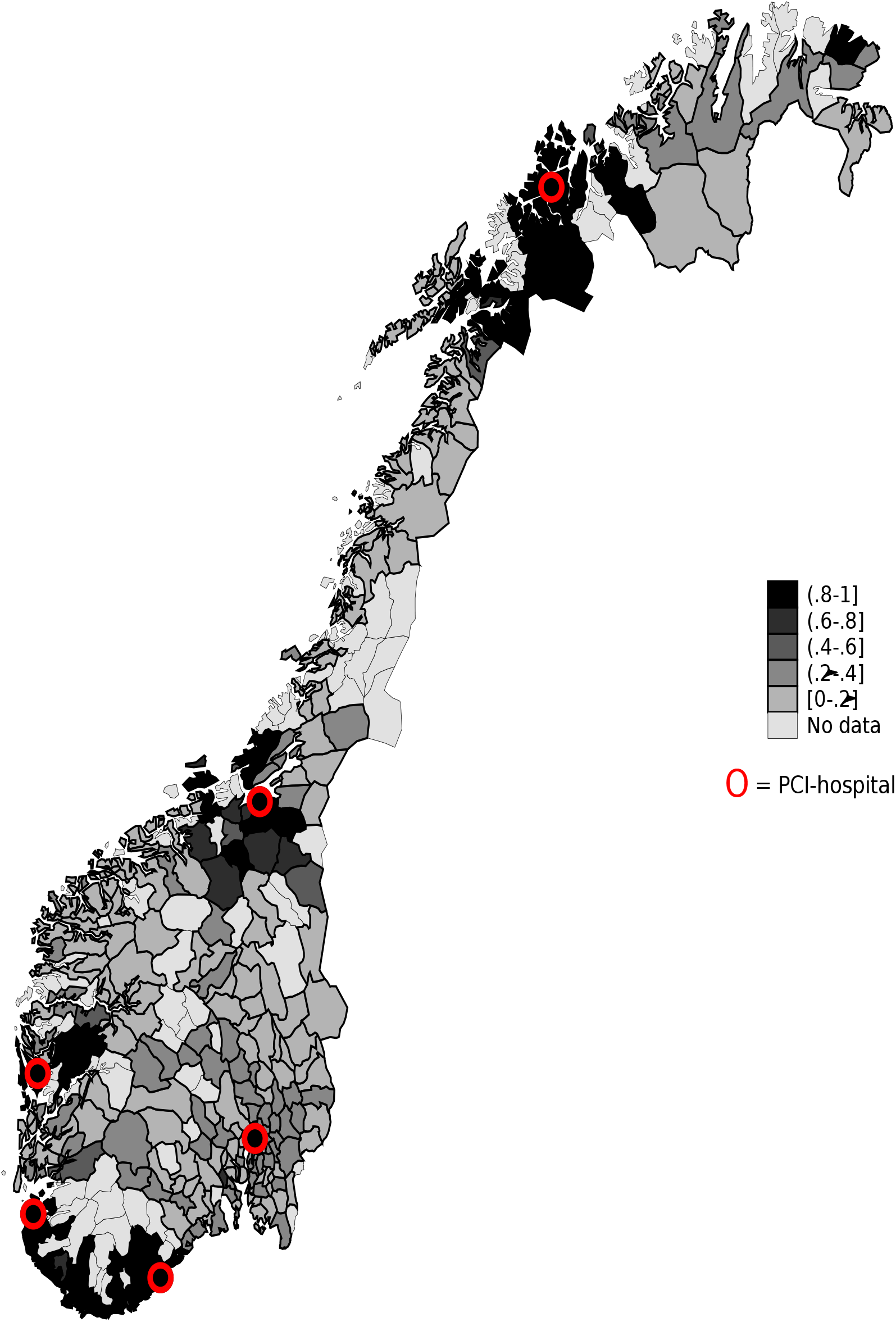
Share of AMI patients send directly to a PCI-hospital in each municipality from 2010-2015. PCI-hospitals location are marked by circles.

## 3 Data and methods

We utilize data from the Norwegian Patient Registry (NPR). NPR contains complete patient level observations (diagnosis, exact date and place for admission and discharges, the degree of urgency at arrival at the health institution) for all somatic public hospitals and private hospitals, which have contracts with regional health authorities. We merge NPR with nationwide individual-level data from Statistics Norway (age, gender, immigration status, municipality and district of resistance, date of death) using the encrypted version of a unique personal identification number issued to every resident of Norway at birth or upon first immigration.

### 3.1 Study sample and treatment variables

Our sample consist of individuals diagnosed with AMI for the first time in at least two years in 2010-2015, using data from 2008-2015. We compare health outcomes for patients diagnosed with AMI who are sent directly to a PCI-hospital with similar patients who are sent to a non-PCI-hospital (and may or may not be transferred to a PCI-hospital). To do this, we first identify all hospitalization spells that overlap with the admission where the patient is diagnosed with AMI as their primary diagnosis (hereafter called AMI spell). Specifically, an AMI spell includes all hospital admissions where there is no more than one day between last discharge and the next admission and where the patient is diagnosed with AMI in at least one of the admissions covered by the spell. We only include AMI spells where the first admission in the spell was registered as acute. A patient is defined as sent directly to a PCI-hospital if the first admission in the spell was to a hospital performing PCI, and directly to a non-PCI-hospital if the first hospital in the spell was a local hospital not performing PCI.

The instrument used is the historic municipal share of acute AMI patients sent directly to a PCI-hospital. For each AMI patient in our sample we calculate the instrument as the historic share of acute AMI patients sent directly to a PCI-hospital in the same municipality (and the 15 city districts in the municipality of Oslo) in the 365 days preceding the current AMI spell of the patient. Since the main sample consists of AMI patients from 2010-2015, we use data from 2009-2015 in this calculation. If there were fewer than ten AMI patients admitted in the municipality the previous year, the given individual is excluded from our sample. This was the case for 8 280 individuals, and leaves us with a sample of 53 773 individuals residing in 338 municipalities.

To compare patients sent directly to a PCI-hospital and directly to a hospital not performing PCI, we link to data on gender, age and date of death as well as municipality (district in Oslo) of resistance at the beginning of the year of AMI spell.

### 3.2 Statistical analyses

First, we show that being sent directly to a PCI-hospital is not random by mapping the share of AMI patients sent directly to a PCI-hospital in each municipality (using the sample of 53 773 patients). Second, to study the effect of initially being sent to a PCI-hospital on mortality we use several multivariate risk adjustment methods as well as the instrumental variable approach. In all models we control for observable characteristics such as age (dummy variable for yearly age groups), gender, and interactions between these, as well as time and municipality fixed effects. Importantly, the municipality fixed effects control for any time-invariant differences between the municipalities, such as absolute and relative distance to different hospitals.

The multivariate risk adjustment methods used to estimate the effect of being sent directly to a PCI-hospital on the probability of death, are linear probability (OLS) and logit models. The implemented model is illustrated by the OLS-equation (1):

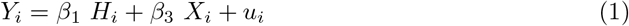

where *Y*_*i*_ is a dummy variable equal 1 if the patient is dead within a set time period, say 1 month, after the start of the AMI spell (otherwise 0). *H*_*i*_ takes the value 1 if individual i is initially sent to a PCI-hospital and 0 of the individual is initially sent to a non PCI-hospital. *X*_*i*_ is a vector of observable characteristics for individual i, including age (dummy variable for yearly age groups), gender,interactions between these. We also include time and municipality fixed effects.

These models identify the effect of being sent directly to a PCI-hospital on a health outcome under the strong assumption that the variation in who are sent directly is (conditionally) uncorrelated with unobserved determinants of the health outcome. However, it is likely that the observable confounders will not capture all differences in health that affect the decision of whether or not to send a patient directly to a PCI-hospital. First, the treatment guidelines are different for different types of AMI. While patient with STEMI heart attack have the most acute condition, they are more often diagnosed in the ambulance, and have higher probability of being sent directly to a PCI-hospital. Since STEMI patients also have higher in-hospital mortality, standard regression results would be expected to be biased towards finding worse health outcomes for those sent directly to a PCI-hospital. On the other hand, while evidence suggests that timely invasive management strategies primarily benefit elderly or high-risk patients, several studies have found that this intervention in practice is directed to lower-risk patients (Pilote et al., 1996). Hence, estimates adjusting for observable characteristics are likely contaminated by omitted variable bias, though the direction of that bias is not clear.

To address concerns of omitted variable bias and endogeneity, we apply the instrumental variable (IV) model which can be illustrates as follows:

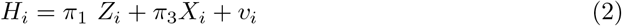

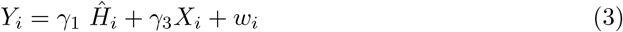

where the instrument *Z*_*i*_ is historic municipal share of patients being sent directly to a PCI-hospital over the preceding year. Hence, since comparing patients with respect to the actual treatment received (sent directly to a PCI-hospital or not) may be biased by health characteristics of the specific patient, our instrumental variable analysis compares groups of patients that differ in the likelihood of being sent directly to PCI-hospital for reasons not related to the health condition of the specific patient. *H* is a dummy variable set to 1 if patient *i* is sent directly to a PCI-hospital, and *Ĥ* is the predicted probability of being sent directly to a PCI-hospital. Note that the instrument takes on the same value only for patients in the same municipality whose AMI spell starts on the same day, and it can thus be different for patients in different municipalities or in the same municipality on different days.

The first-stage regression in equation 2 estimates to what degree the instrument affects the probability of being sent directly to a PCI-hospital. The second stage regression in equation 3 provides the main parameter of interest, *γ*_1_, which captures the effect of being sent directly to a PCI-hospital, instead of first going to a non-PCI-hospital, for patients for whom the hospital they are sent to (PCI or not) shifts as a result of variation in the practice of the municipality (i.e. in the instrument *Z*_*i*_) (i.e. the local average treatment effect, LATE). Like in the multivariate risk adjustment models, we control for a vector of observable characteristics (*X*_*i*_) and time and municipality fixed effects. We estimate the LATE using two-stage least square (2SLS), and cluster on municipality level.

In order for the identification strategy to be valid, the independence assumption must hold, meaning that our instrument should be uncorrelated with individual patients’ potential outcomes. If this assumption holds, the intention to treat estimates (ITT) of the effect of the instrument on the outcome will estimate causal effects of being sent directly to a PCI-hospital vs. first to a non-PCI-hospital. This seems reasonable in our situation, as the share of preceding patients who are sent to a PCI-hospital is not likely to be determined by characteristics of future individual patients, which is also suggested empirically: Table A1 shows that, as expected, observable characteristics of the patients are predictive of actually being sent directly to a PCI-hospital, but it also shows that the observable characteristics of the patients are not predictive of previous patients in the municipality being sent directly to a PCI-hospital (i.e. the instrument).^1^

To identify the local average treatment effect (LATE), the so-called exclusion restriction must hold. It requires that the instrument only affects the outcome of interest through the treatment. Put differently, the health outcome (mortality) of a given patient should not be directly affected by the share of previous patients admitted directly to a PCI-hospital. Congestion, where the current patient is sent to a non-PCI-hospital because the previous patients were sent to a PCI-hospital, would lead to a violation of this assumption. To take this into account we do not include admissions at the same day as the given patient when we calculate the instrument (i.e., we calculate a leave-out-mean). Also the instrument must be highly correlated with the probability of being sent directly to a PCI-hospital, which we can easily confirm empirically in the first stage in equation 2. The monotonicity assumption, requires that the instrument should (weakly) change the probability of treatment in the same direction. In our setting, we need to assume that a higher rate of patients being sent directly to a PCI-hospital (weakly) increases the likelihood of future patients being sent directly to a PCI-hospital. A possible violation of this assumption occurs if healthcare professionals experience that the health outcomes for patients sent directly is poor and therefore send fewer patients directly to a PCI-hospital in the future. However, we argue this is unlikely as emergency medical personnel often do not observe the health outcome of the patient.

If these assumptions hold the estimate of *γ*_1_ captures the local average treatment effect (LATE) of being sent directly to a PCI-hospital for the compliers. In our setting with municipality fixed effects, compliers are patients who are sent directly to a PCI-hospital because there is a *change over time* in the municipality’s inclination of sending patients directly to a PCI-hospital.

Analyses were performed by using STATA 16.

## 4 Results

Figure 1 shows a map over the share of AMI patients that is sent directly to a PCI-hospital in each municipality in Norway 2010-2015. There is substantial variation in the share initially admitted to a PCI-hospital. For obvious reasons, individuals living in municipalities close to a PCI-hospital are more often sent directly than those living in municipalities further away.

Table 1 gives the summary statistics for the full sample (N=53 773), where 38 percent are sent directly to a PCI-hospital (N=20 336) and the remaining 62 percent are sent first to a non-PCI-hospital (N=33 437). Of those who were sent first to a non-PCI-hospital, 70.4 percent is forwarded to a PCI-hospital in the same AMI spell. Those sent directly to a PCI-hospital are more often male, younger, and are less likely to have comorbidities than those sent first to a non-PCI-hospital. They are also less likely to die during the first month compared to those sent to a non-PCI-hospital (7.5 % vs. 8.4 %).

**Table 1:**
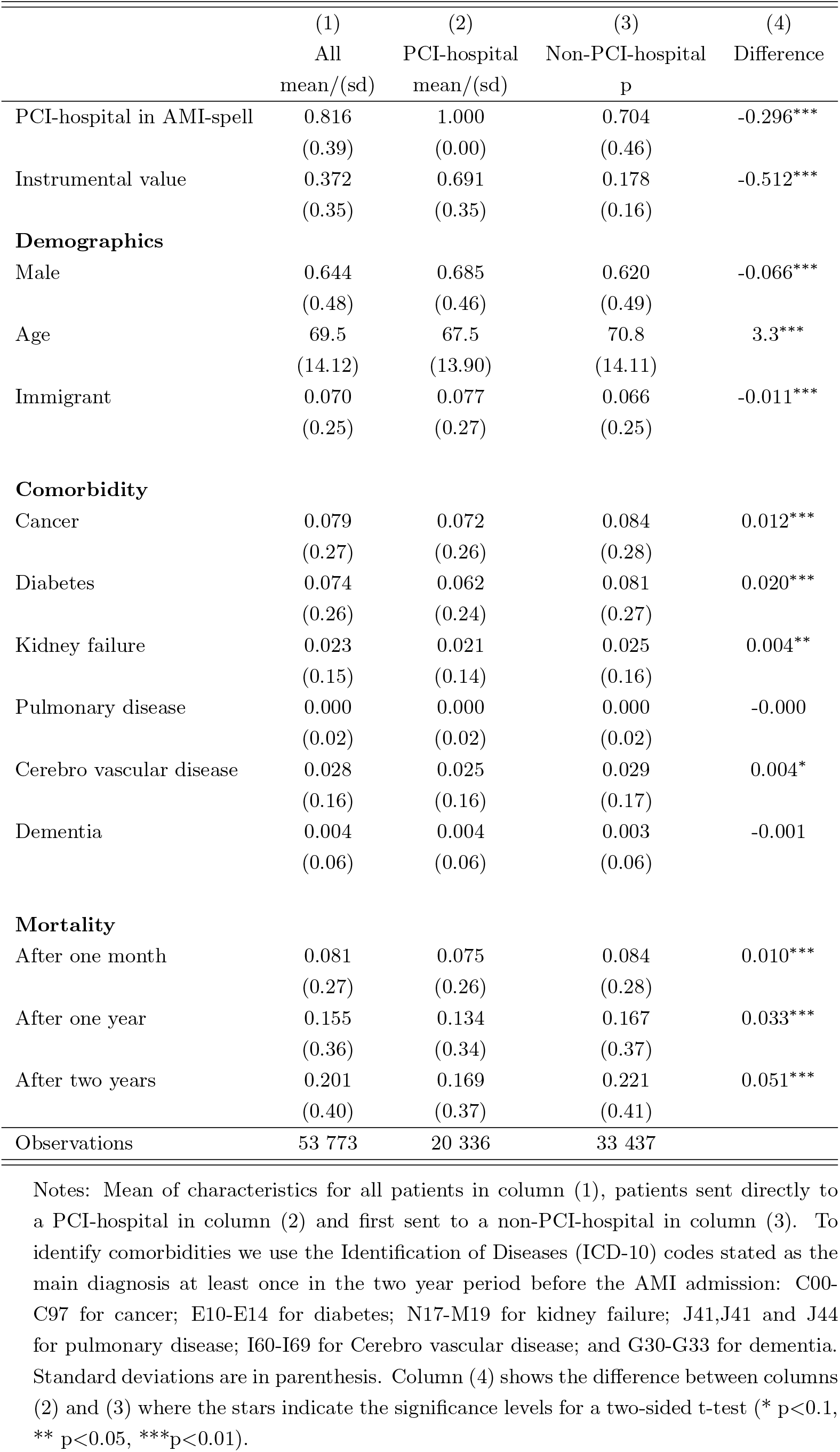
Mean charactersitics of all patients in the sample, and of patients first sent to a PCI-hospital vs. to a non-PCI-hospital

|  | (1) | (2) | (3) | (4) |
| --- | --- | --- | --- | --- |
|  | All | PCI-hospital | Non-PCI-hospital | Difference |
|  | mean/(sd) | mean/(sd) | p |  |
| PCI-hospital in AMI-spell | 0.816<br>(0.39) | 1.000<br>(0.00) | 0.704<br>(0.46) | -0.296*** |
| Instrumental value | 0.372<br>(0.35) | 0.691<br>(0.35) | 0.178<br>(0.16) | -0.512*** |
| **Demographics** |  |  |  |  |
| Male | 0.644<br>(0.48) | 0.685<br>(0.46) | 0.620<br>(0.49) | -0.066*** |
| Age | 69.5<br>(14.12) | 67.5<br>(13.90) | 70.8<br>(14.11) | 3.3*** |
| Immigrant | 0.070<br>(0.25) | 0.077<br>(0.27) | 0.066<br>(0.25) | -0.011*** |
| **Comorbidity** |  |  |  |  |
| Cancer | 0.079<br>(0.27) | 0.072<br>(0.26) | 0.084<br>(0.28) | 0.012*** |
| Diabetes | 0.074<br>(0.26) | 0.062<br>(0.24) | 0.081<br>(0.27) | 0.020*** |
| Kidney failure | 0.023<br>(0.15) | 0.021<br>(0.14) | 0.025<br>(0.16) | 0.004** |
| Pulmonary disease | 0.000<br>(0.02) | 0.000<br>(0.02) | 0.000<br>(0.02) | -0.000 |
| Cerebro vascular disease | 0.028<br>(0.16) | 0.025<br>(0.16) | 0.029<br>(0.17) | 0.004* |
| Dementia | 0.004<br>(0.06) | 0.004<br>(0.06) | 0.003<br>(0.06) | -0.001 |
| **Mortality** |  |  |  |  |
| After one month | 0.081<br>(0.27) | 0.075<br>(0.26) | 0.084<br>(0.28) | 0.010*** |
| After one year | 0.155<br>(0.36) | 0.134<br>(0.34) | 0.167<br>(0.37) | 0.033*** |
| After two years | 0.201<br>(0.40) | 0.169<br>(0.37) | 0.221<br>(0.41) | 0.051*** |
| Observations | 53 773 | 20 336 | 33 437 |  |

Notes: Mean of characteristics for all patients in column (1), patients sent directly to a PCI-hospital in column (2) and first sent to a non-PCI-hospital in column (3). To identify comorbidities we use the Identification of Diseases (ICD-10) codes stated as the main diagnosis at least once in the two year period before the AMI admission: C00-C97 for cancer; E10-E14 for diabetes; N17-M19 for kidney failure; J41, J41 and J44 for pulmonary disease; I60-I69 for Cerebro vascular disease; and G30-G33 for dementia. Standard deviations are in parenthesis. Column (4) shows the difference between columns (2) and (3) where the stars indicate the significance levels for a two-sided t-test (\* $p < 0.1$ , \*\* $p < 0.05$ , \*\*\* $p < 0.01$ ).

### 4.1 Multivariate risk adjustment methods

The OLS results show that AMI patients who are sent directly to a PCI-hospital have 1.4 percentage points higher probability of death during the first month after the AMI spell than patients who are sent first to a non-PCI-hospital (Figure 2). Unlike the short-time results, the long-term (24 month) mortality results indicate that those sent directly to a PCI-hospital have 0.8 percentage points lower mortality.^2^ As elaborated on above, health care personnel have to make complex treatment decisions in a short period of time, and are likely to base these decisions on circumstances of the acute health event that are unobservable to reseachers. To circumvent such selection at the individual level, we instrument whether the current patient is sent directly with the historical share of patients in the same municipality who are sent directly to a PCI-hospital.

**Figure 2:**
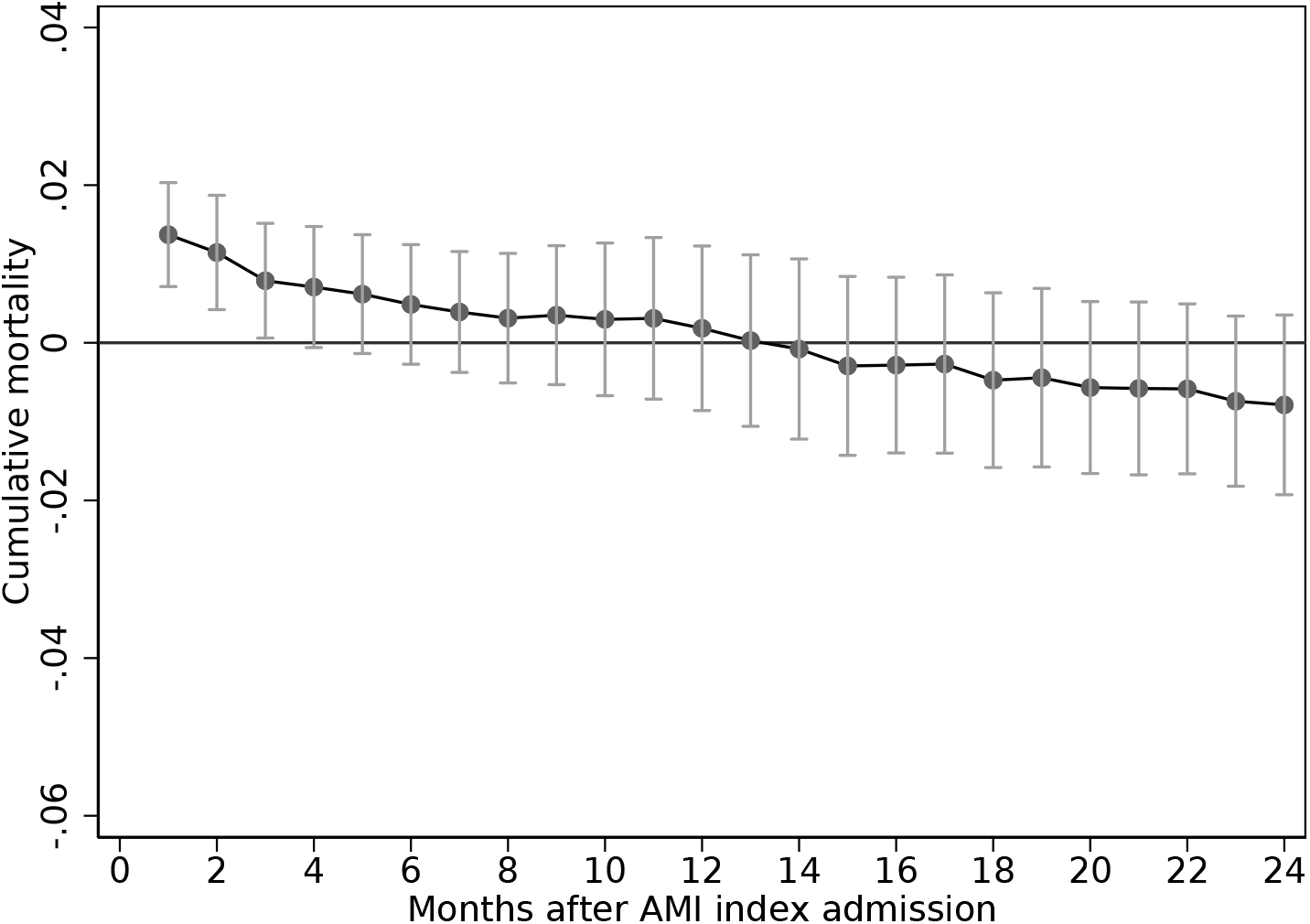
The association between being sent directly to a PCI-hospital and mortality (OLS) Notes: The figure show OLS associations (95 % confidence interval) between being sent directly to a PCI-hospital vs to a non-PCI-hospital and mortality in different time intervals. More details on model specification is found in A2.

### 4.2 Instrumental variable

The instrument predicts clearly significantly who are sent directly to a PCI-hospital (F-statistics of 12; see Table A2). Patients in municipalities with historic higher share of patients sent directly to a PCI-hospital have lower mortality than comparable patients in municipalities where fewer have been sent directly to a PCI-hospital (i.e. the ITT-estimates, from our instrumental variable approach are negative for all time intervals). This suggests that a marginal increase in the share sent directly to a PCI-hospital would reduce mortality (Table 2, Figure 3). The results are, however, not statistically significant. Scaling the ITT estimates by the first-stage estimates provides the LATE, and it shows that those sent directly to a PCI-hospital have 4.8 percentage points decrease (95% CI - 18.1 to 8.5) in mortality after 1 month compared to those sent first to a non-PCI-hospital. These patients who are moved into treatment due to increased municipal inclination to send patients directly to a PCI-hospital, have a lower mortality at all time intervals (Table 2, Figure A4)^3^. However, none of the results are statistically significant.

**Table 2:** The association between the historic share sent directly to PCI-hospital on mortailty at indicated time intervals (ITT) and the effect of being sent directly to a PCI-hospital on mortality (LATE)

|  | (1) | (2) | (3) |
| --- | --- | --- | --- |
|  | 1 month | 12 months | 24 months |
|  | b/se | b/se | b/se |
| ITT (OLS) | -0.012<br>(0.016) | -0.008<br>(0.019) | -0.026<br>(0.021) |
| LATE (2SLS) | -0.048<br>(0.068) | -0.034<br>(0.078) | -0.105<br>(0.093) |
| FS (OLS) | 0.247***<br>(0.071) | 0.247***<br>(0.071) | 0.247***<br>(0.071) |
| F-stat | 12 | 12 | 12 |
| *N* | 53773 | 53773 | 53773 |
| ITT (Logit) | -0.012<br>(0.016) | -0.009<br>(0.020) | -0.027<br>(0.021) |
| *N* | 52827 | 53241 | 53443 |

Note: This table reports the Intention to treat (ITT), the local average treatment effect (LATE) and the first stage (FS) results for the OLS regression, as well as the ITT result for the Logit regression. The table reports differences in mortality rates after 1, 12 and 24 months from OLS and Logit models. All regressions control for age, gender, an interaction between these, year and municipality fixed effect. Regression results from the Logit model are reported as the average marginal effect (AME). Standard errors are clustered at municipality level. Stars indicate significance levels (\* $p < 0.1$ , \*\* $p < 0.05$ , \*\*\* $p < 0.01$ )

**Figure 3:**
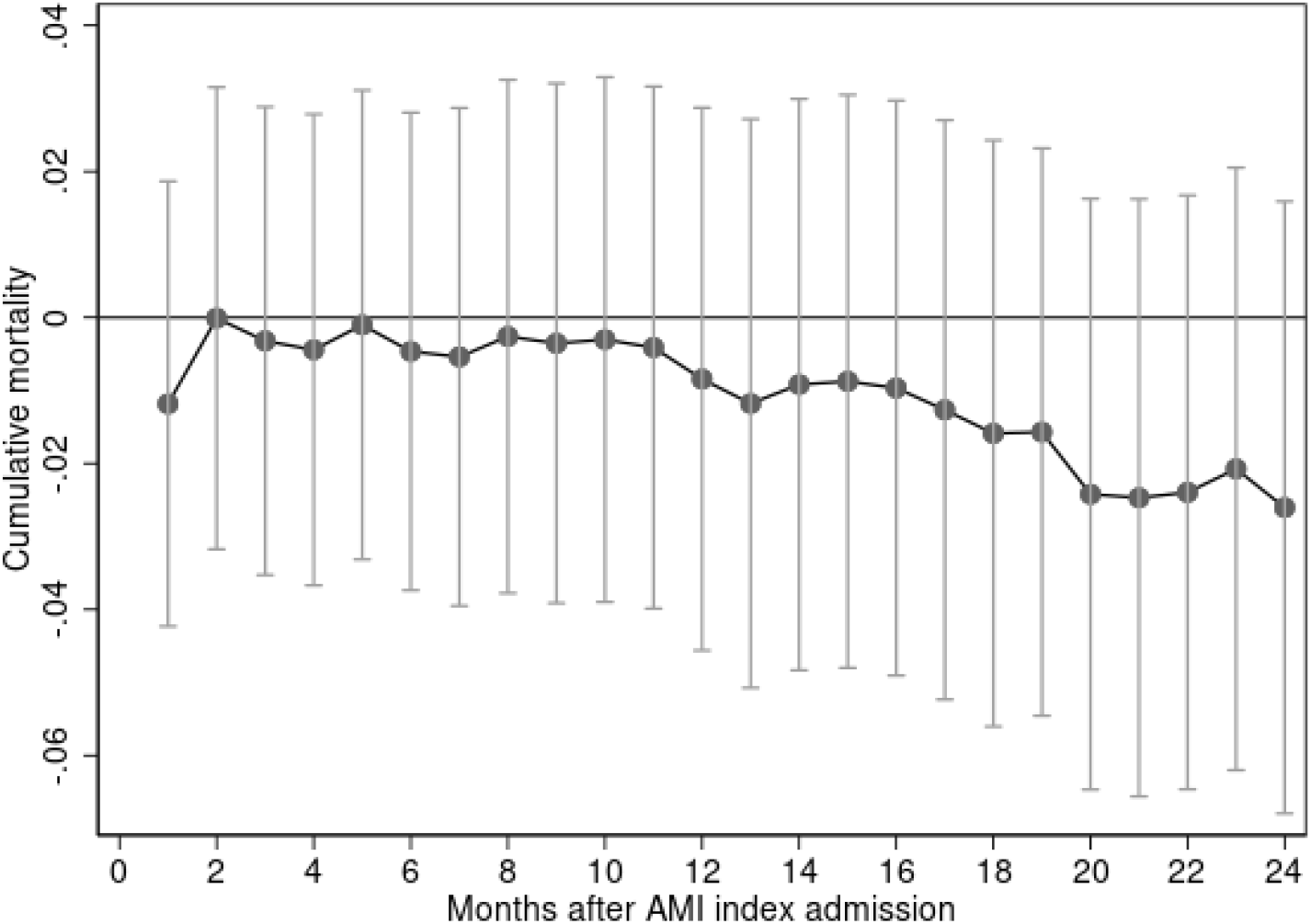
The association between historic share sent directly to a PCI-hospital and morality (ITT) The figure show associations (95% confidence interval) between the historic share sent directly to a PCI-hospital in the municipality and the mortality of the current patient, in different time intervals. More details on model specification is found in Table 2.

## 5 Discussion

We show vast geographic variation in the share of AMI patients sent directly to a PCI-hospital, and that being sent to a PCI-hospital is highly correlated with observable characteristics of the patient. Using the instrument to handle selection by observable and non-observable characteristics, estimates of the local average treatment effect (LATE) show that those sent directly to a PCI-hospital have 4.8 percentage points (95% CI - 18.1 to 8.5) lower mortality after 1 month than patients sent first to a non-PCI-hospital. However, the estimates are too imprecise to conclude that health personnel should change their practice and send more patients directly to a PCI-hospital.

This empirical study emphasises the importance of carefully considering possible selection issues using observational data to shed light on possible changes in health personnels’ propensity to assign AMI patients to local hospitals or hospitals with PCI farther away. Our study my thus contribute to existing literature on the best treatment strategies for AMI patients. A national cohort study evaluating 158 831 elderly Medicare patients hospitalized with AMI in the U.S found that invasive medical treatment, such as thrombolysis, reduced the incremental benefit of more expensive treatments such as invasive surgery (Stukel et al., 2005). In addition, a randomized controlled trial of 1 653 individuals found that thrombolyisis became superior to PCI when the PCI-related delay is prolonged and exceeded the guideline-mandated times (Gershlick et al., 2015). In line with these studies our results indicate that there is only a small and not statistically significant decrease in mortality associated with marginal increases in the share sent directly to a PCI-hospital. Our findings may therefore imply that health personnel navigate the patients to the best treatment option based on factors such as travel time and severity of the disease.

An important strength of our study is the use of nation-wide registry data that included information on all hospital admissions in Norway. Additionally, we used an instrumental variables approach to handle non-random selection of which patients are sent directly to a PCI-hospital or not. However, there are also certain limitations to our analysis. First, our retrospective study including individuals diagnosed with AMI at a hospital during the study period, and we did not have data on care given before entering the hospital. Hence, we do not know whether the health personnel were able to diagnose the patient in the ambulance or whether the patient was given any pre-hospital treatment such as trombolysis. Second, our instrument is not very strong (F-stat of 12), which may cause biased and imprecise estimates. Further research should strive to find stronger instruments or use other empirical methods to estimate the effect of being initially sent to a PCI-hospital. Third, for the identification strategy to be valid the independence assumption must hold, meaning that the historic local share sent directly to a PCI-hospital (the instrument) should be uncorrelated with the current patient’s observed and unobserved pre-admission characteristic. We have argued that this is likely to hold as the share of preceding patients who are sent to a PCI-hospital is not likely to be determined by characteristics of the current patient. However, if there are trends within municipalities that change over time, that is correlated with the probability of being sent directly and correlated with health outcomes, this may not hold. Since it is not possible to provide a definite answer to the validity of this assumption, caution must be taken when interpreting results from this observational study too.

We have only estimated the average effect at the national level, and the effect of increasing the share of patients sent directly to a PCI-hospitals may differ between local areas with different absolute and relative transportation times to non-PCI- and PCI-hospitals. It may thus be possible that local guidelines on how to treat AMI patients could be improved, but this is hard to evaluate without well-crafted randomized controlled trails.

## 6 Conclusion

Identifying health outcomes of sending patients directly to a PCI-hospital is difficult because of non-random selection. To handle selection bias we used an instrumental variable approach with historic shares of previous AMI patients sent directly to a PCI-hospital as an instrument for the propensity of sending the next AMI patient directly to a PCI-hospital. While our OLS results show that AMI patients sent directly to a PCI-hospital have higher short term mortality, our instrument variable approach indicates the opposite: being sent directly to a PCI-hospital reduces short term survival, but results are statistically nonsignificant. Thus, the results from traditional multivariable risk adjustment methods may be taken to suggest that more AMI-patients should be admitted to the local non-PCI-hospital, while the instrumental variable results may suggest that health personnel navigates patients to the hospital that can give the appropriate treatment. However, the estimates are too imprecise to conclude that health personnel should change their practice.

## Data Availability

The datasets that support the findings of this study contain sensitive information and are not publicly available due to privacy laws. Individual-level data for research are generally available within Norway upon application conforming with strict regulations and procedures.

## Declarations

### Ethics statement

Permission to use patient data from Norwegian Regional Ethics Committee was given(#2017/373).

### Founding

This paper has received funding from the Research Council of Norway (grant #256678 and #256678).

### Author contributions

All authors designed the study. MG performed the statistical analyes and drafted the manuscript. KET and HØ critically evaluated and critically revised all stages of the research process. All authors gave final approval for the version to be submitted.

## Acknowledgements

We would like to thank Ingrid Huitfeldt and Simon Bensnes for helpful discussions, suggestions and comments at various stages of this project. The interpretation and reporting of the data are the sole responsibility of the authors.

## A Appendix

**Table A1:** Instrument validity

|  | (1) | (2) |
| --- | --- | --- |
|  | Initially to PCI-hospital | Instrumental value |
|  | b/se | b/se |
| Age | -0.003***<br>(0.000) | -0.000<br>(0.000) |
| Male | 0.031***<br>(0.004) | -0.001<br>(0.001) |
| Immigrant | -0.008<br>(0.007) | 0.001<br>(0.001) |
| Cancer | -0.012*<br>(0.005) | -0.002**<br>(0.001) |
| Diabetes | -0.037***<br>(0.006) | 0.001<br>(0.001) |
| Kidney failure | -0.012<br>(0.012) | -0.001<br>(0.002) |
| Pulmonary disease | -0.092<br>(0.052) | 0.011<br>(0.009) |
| Cerebro vascular disease | -0.024***<br>(0.007) | 0.000<br>(0.002) |
| Dementia | 0.006<br>(0.030) | 0.000<br>(0.003) |
| N | 53773 | 53773 |
| Dep. mean | 0.378 | 0.372 |
| Joint F-statistics | 19.75 | 41.17 |
| Joint P-value | 0.000 | 0.3158 |

Note: Table shows estimates of actually being sent directly to a PCI-hospital vs. first being sent to a non-PCI-hospital (column 1) and the historic share of patients in the municipality of the patient being sent directly to a PCI-hospital vs first being sent to a non-PCI-hospital (i.e. the instrument) (column 2) on observable patient characteristics measured prior to the start of the AMI spell. Both models include fixed effects for year and municipality, and are clustered on municipality. Stars indicate significance level (\* $p < 0.1$ , \*\* $p < 0.05$ , \*\*\* $p < 0.01$ )

**Table A2:**
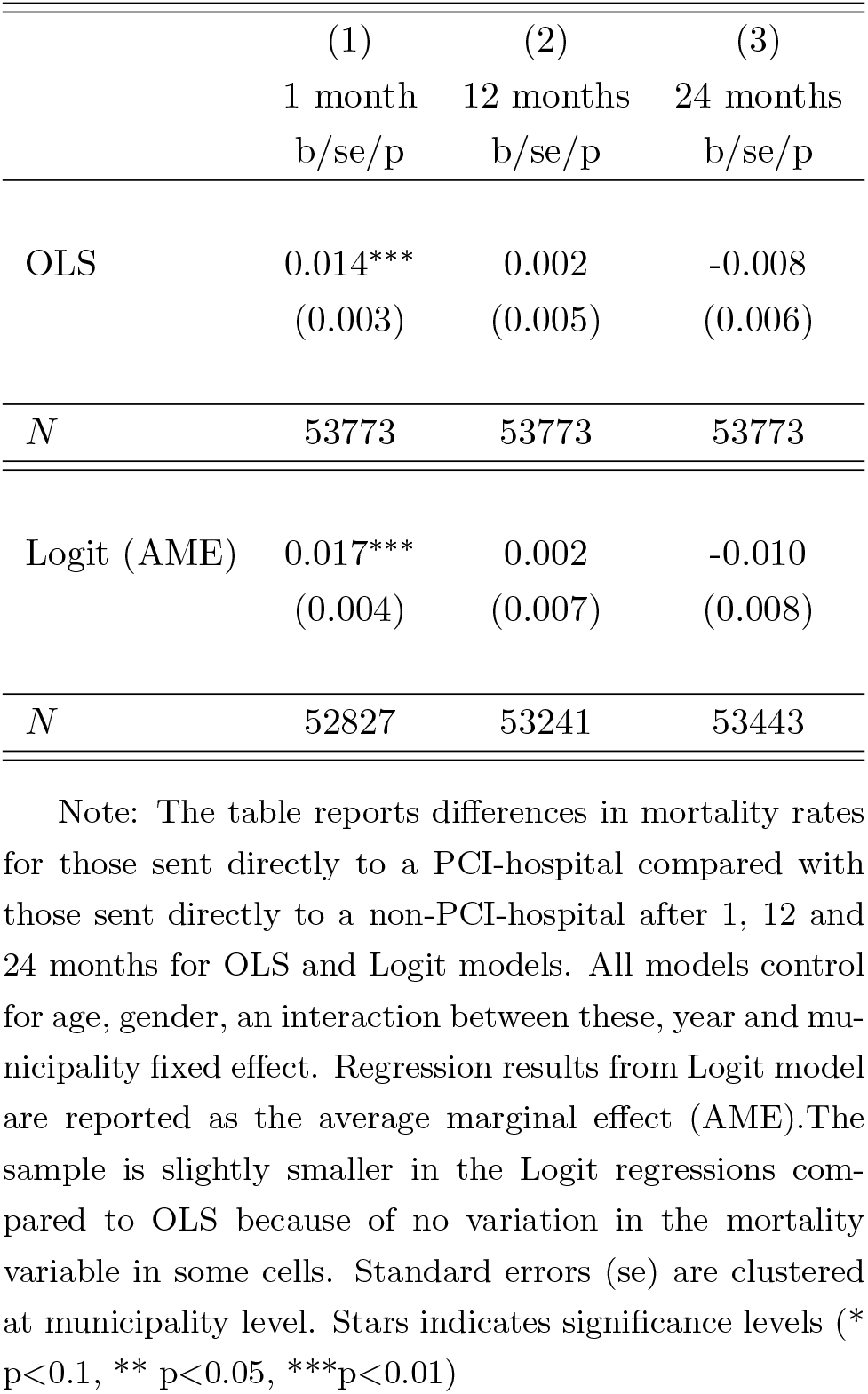
The association between initial admission to a PCI-hospital vs. non-PCI-hospital on mortality at indicated time intervals

|  | (1) | (2) | (3) |
| --- | --- | --- | --- |
|  | 1 month | 12 months | 24 months |
|  | b/se/p | b/se/p | b/se/p |
| OLS | 0.014***<br>(0.003) | 0.002<br>(0.005) | -0.008<br>(0.006) |
| *N* | 53773 | 53773 | 53773 |
| Logit (AME) | 0.017***<br>(0.004) | 0.002<br>(0.007) | -0.010<br>(0.008) |
| *N* | 52827 | 53241 | 53443 |

Note: The table reports differences in mortality rates for those sent directly to a PCI-hospital compared with those sent directly to a non-PCI-hospital after 1, 12 and 24 months for OLS and Logit models. All models control for age, gender, an interaction between these, year and municipality fixed effect. Regression results from Logit model are reported as the average marginal effect (AME). The sample is slightly smaller in the Logit regressions compared to OLS because of no variation in the mortality variable in some cells. Standard errors (se) are clustered at municipality level. Stars indicates significance levels (\* $p < 0.1$ , \*\* $p < 0.05$ , \*\*\* $p < 0.01$ )

**Figure A4:**
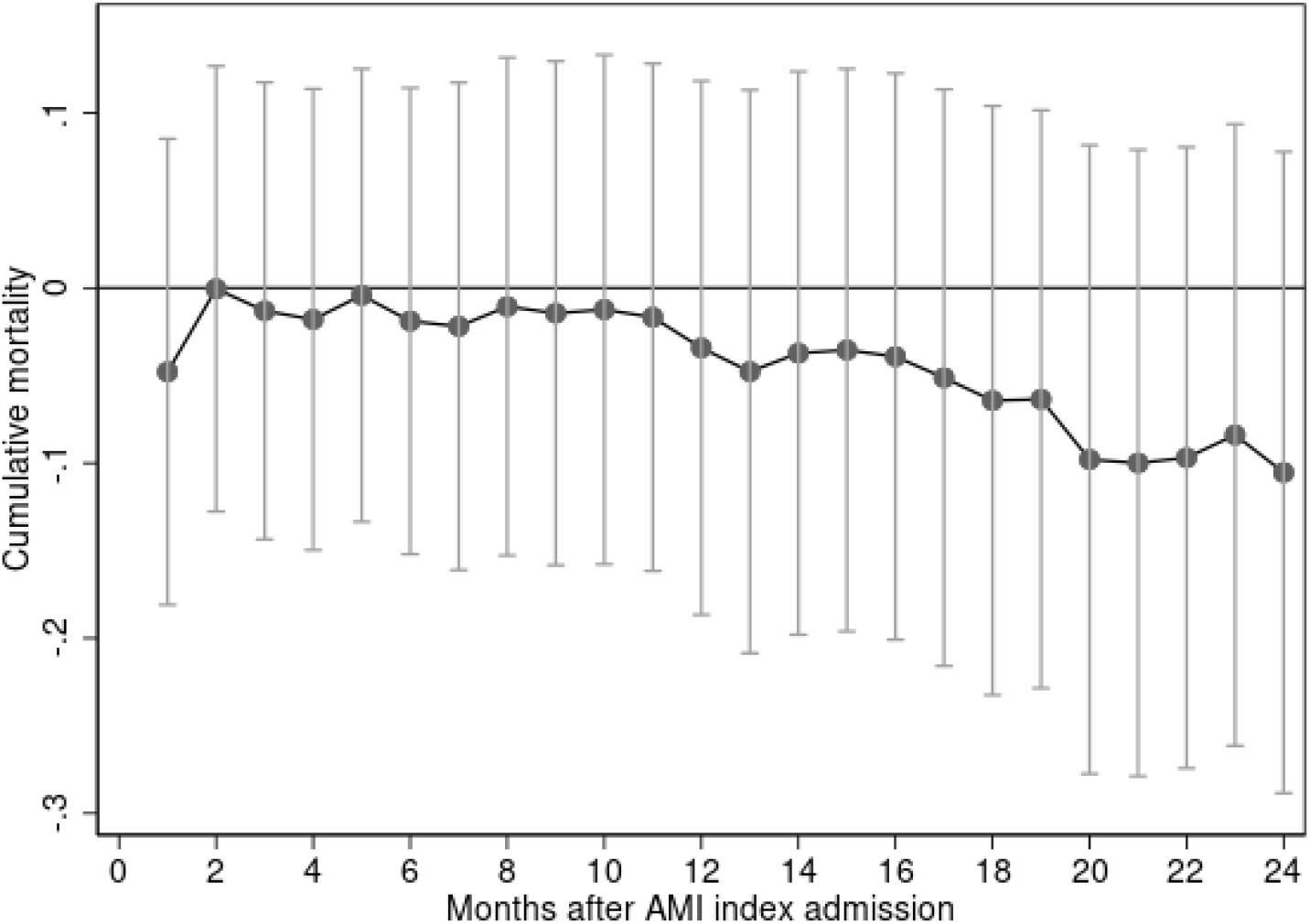
The association between historic share sent directly to a PCI hospital and morality (2SLS) The figure show associations (95% confidence interval) between the historic share sent directly to a PCI-hospital in a municipality and the mortality of the current patient for the compliers, in different time intervals. More details on model specification is found in Table 2.

**Table A3:** The effects of initial admission to PCI-hospital on Mortality at indicated time intervals after Acute Heart Attack, alternative instrument

|  | (1) | (3) | (5) |
| --- | --- | --- | --- |
|  | 1 month | 12 months | 24 months |
|  | b/(se) | b/(se) | b/(se) |
| ITT(OLS) | -0.003<br>(0.009) | 0.001<br>(0.012) | -0.005<br>(0.012) |
| IV (2SLS) | -0.020<br>(0.072) | 0.004<br>(0.098) | -0.041<br>(0.096) |
| FS | 0.127***<br>(0.032) | 0.127***<br>(0.032) | 0.127***<br>(0.032) |
| F-test | 15 | 15 | 15 |
| *N* | 59913 | 59913 | 59913 |

Note: The alternative instrument equals the share of the ten last AMI patient in a local area that was sent directly to a PCI-hospital. All regressions include controls for age, gender and their interactions, as well as municipality and time fixed effects. Standard errors are clustered at municipality level. Stars indicates significance levels (\* $p < 0.1$ , \*\* $p < 0.05$ , \*\*\* $p < 0.01$ )

## Footnotes

1 There is one covariate that is significantly predicting the instrument the 5% level; cancer. Given the number of covariates being tested, the probability of getting one boarder line significant variable by pure chance is high. We also see that patient characteristics are jointly correlated with actual treatment but not with the instrument (jointly p-values is 0.00 for the actual treatment and 0.32 for the instrument).

2 Results from Logit models are very similar; see Table where results are reported as average marginal effects for the Logit model too to enable comparison).

3 While the instrument in our main analysis is constructed from the historic share sent directly to a PCI-hospital (or not) over the last year, we have also run a robustness check with an alternative instrument calculated as the share sent directly to PCI-hospital based on the ten last AMI patient in each municipality. The results are shown in Table A3 and they are largely in line with those reported in our main analysis.

